# Facilitators and barriers when implementing antibiotic stewardship interventions in neonates at risk of early-onset sepsis

**DOI:** 10.1101/2025.03.31.25324838

**Authors:** Liesanne E.J. van Veen, Sanne W.C.M. Janssen, Gerdien A. Tramper-Stranders, Niek B. Achten, Annemarie M.C. van Rossum, Frans B. Plotz, Erwin Ista

**Affiliations:** Department of Pediatrics, Franciscus Gasthuis & Vlietland, Rotterdam, The Netherlands; Department of Paediatrics, Erasmus MC, Sophia Children’s Hospital, Rotterdam, The Netherlands; Department of Paediatrics, Tergooi MC, Hilversum, The Netherlands; Department of Paediatrics, Amsterdam UMC, Amsterdam, The Netherlands; Department of Internal Medicine, Division of Nursing Science, Erasmus MC University Medical Center Rotterdam, Rotterdam, The Netherlands; Department of Neonatal and Pediatric Intensive Care, Division of Pediatric Intensive Care, Erasmus MC-Sophia Children’s Hospital, Rotterdam, The Netherlands

## Abstract

**Introduction:** Antibiotic stewardship is becoming increasingly important in neonatal care, given the impact of early antibiotic use on hospitalisation, short– and long-term health, and antibiotic resistance. To tackle these challenges, various antibiotic stewardship interventions have been developed. While neonatal early-onset sepsis (EOS) interventions have demonstrated potential to reduce and optimize antibiotic use, evidence regarding their implementation remains limited. In this study we aimed to identify barriers and facilitators of implementing three evidence-based antibiotic stewardship interventions in EOS care.

**Methods:** Interdisciplinary focus group interviews were conducted with paediatricians, neonatal nurses, paediatric residents, midwives, primary care maternity nurses, microbiologists, pharmacists, and general practitioners. A semi-structured interview guide was used to discuss the EOS calculator, PCT-guided therapy, IV-oral switch therapy and current EOS care practices. The Consolidated Framework for Implementation Research (CFIR), consisting of 5 determinant domains (innovation, outer setting, inner setting, individuals’, and implementation process domain), was used to guide the interviews and data analyses, using a rapid deductive content analysis approach.

**Results:** Eleven focus group interviews were conducted with 81 participants. We identified 34 barriers and 20 facilitators. Most barriers concerned the inner setting (n=11), intervention characteristics (n=10), and the individual health professional level (n=8), while most facilitators were related to the intervention characteristics (n=8). Overarching barriers for implementing novel antibiotic stewardship interventions were external pressure to adhere to the national guidelines or affiliated academic regional protocols and the expected care shift towards healthcare workers with an already high workload. Universal dissatisfaction with the current national guideline and a hospital culture of evidence-based, patient-centred care in the presence of a strong opinion leader were reported as facilitators of implementing antibiotic stewardship interventions.

**Conclusion:** This study identified barriers and facilitators influencing the implementation of three antibiotic stewardship interventions in neonatal EOS care. These determinants can be consolidated in the following themes: balancing quality of evidence with professional core values, managing care shifts and enhancing interdisciplinary communication, and conflicts with un-updated guidelines.

## INTRODUCTION

In high-income countries, 1.2% to 12.5% of liveborn neonates is treated for suspected early-onset sepsis (EOS), which is up to 58 times as much as the incidence ^1^. In the Netherlands, a study across 15 hospitals reported a treatment rate of 4.6% ^2^. However, as there is growing awareness of the negative impact of early antibiotic use on hospitalization rates, short– and long-term health, and antibiotic resistance, antibiotic stewardship is becoming increasingly important ^3, 4^. Clinicians seek strategies to prescribe more cautiously, which is portrayed in the low adherence to the current Dutch national guideline, which still recommends the categorical risk factor assessment approach that results in a low threshold to treat ^5^. The low guideline adherence is accompanied by large practice variation between hospitals, underscoring the need for uniform tools and strategies that help clinicians take the lead in antibiotic stewardship and minimize unnecessary treatment and hospitalisation ^2^.

In response, various interventions have been studied in neonates at risk for EOS, such as serial clinical examinations, the EOS calculator, procalcitonin (PCT)-guided therapy, de-implementation of routine CRP, and IV-to-oral switch therapy ^6–12^. These interventions can either prevent initiation of treatment, shorten the duration, or reduce the invasiveness, and promote family-centred care. In the PrOTeCt-NEO (Promoting Optimal Treatment Choices in Neonates with suspected Early-Onset sepsis) implementation project (NCT06845332), three high-level evidence interventions covering different aspects of EOS management will be implemented and evaluated in Dutch neonatal care. The included interventions are the neonatal EOS calculator, PCT-guided therapy, and the IV-to-oral switch therapy. The EOS calculator, a risk prediction tool that calculates an individual EOS risk based on maternal risk factors and neonatal physical examination, reduced antibiotic treatment initiation by 44% in retro– and prospective observational studies ^6, 8, 9^. The PCT-guided therapy, based on the NeoPInS study algorithm, reduces the length of antibiotic treatment on average of 10 hours in neonates at low and intermediate risk of infection, using two consecutive PCT values below the threshold to confirm absence of infection ^11^. Thirdly, the IV-to-oral switch therapy was found to be effective and safe in neonates in the RAIN study, allowing continuation of treatment at home ^7^.

Evidence suggests that implementing these interventions would safely reduce antibiotics, promote family-centred care, and shorten hospitalization. However, providing convincing evidence and incorporating recommendations into a guideline does not guarantee their application in practice, as guideline non-adherence rates vary from 8.2 to 65.3% ^13–15^. Non-adherence might result in lower quality of care, waste of resources, and variation in provided healthcare ^16, 17^. To achieve sustainable use of novel interventions, it is essential to gather type 3 evidence on dissemination and implementation within context by involving relevant stakeholders ^18^. Understanding stakeholder perspectives will allow for intervention optimalization and actively changing the context in which they are used ^18, 19^. Therefore, we aimed to determine stakeholders’ barriers and facilitators of implementing the EOS calculator, PCT-guided therapy and IV-to-oral switch therapy into neonatal care, to inform future implementation strategies.

## METHODS

### Design

This study adopted a qualitative, descriptive design, using semi-structured focus group interviews among stakeholders in EOS care. This method was chosen as it facilitates efficiently gathering diverse viewpoints and enables group interaction, promoting enrichment of participant’s responses ^20, 21^. This was considered useful for assessing the implementation of three interventions covering multiple professions in neonatal care. The Consolidated Citeria for Reporting Qualitative Research (COREQ) checklist was followed to report this research (Supplemental file 1) ^22^.

### Setting and participant selection

In the Netherlands, neonates at risk of EOS are cared for in neonatal (Level II-IV) and obstetric wards (Level I) ^23^. Paediatricians and paediatric residents evaluate neonates, order laboratory tests, and prescribe antibiotics in consultation with microbiologists and pharmacists if required. Neonatal nurses monitor the neonates and administer antibiotics, either intravenously or orally. Pre– and post-hospital care is provided by midwives, maternity nurses, and general practitioners. Midwives are medically responsible for mother and neonate at home during the first 10 days postpartum with the support of maternity nurses in the home setting. Afterwards, care responsibility transitions to the general practitioner. Within the Protect-Neo implementation project, 11 Dutch secondary hospitals and their first-line neonatal care networks were included, representing regional differences in care and served as primary recruitment sites for participants. Local paediatricians leveraged their networks via phone, email, or in-person communication to suggest potential participants, including paediatricians, residents, nurses, pharmacists, microbiologists, and general practitioners. Obstetric partnerships helped reach midwives and maternity nurses. Participants who agreed to share contact details received study information by one of the researchers via email. Quota sampling was utilized to ensure maximum heterogeneity by aiming to include one participant from each profession per focus group ^24^.

### Guiding Framework

Data collection and analysis were based on the updated Consolidated Framework for Implementation Research (CFIR) ^25^. This framework includes 48 constructs across five domains: the intervention (three antibiotic stewardship interventions), the inner setting (local neonatal care networks; hospital and primary neonatal care), the outer setting (affiliated academic hospitals, national and international level), individuals (primary and secondary care healthcare workers), and the implementation process (activities to implement the three interventions) (Figure 1).

### Data collection

A focus group with the multidisciplinary team as described above was conducted at each of the 11 sites. Additional sessions were held as necessary to achieve data saturation, defined as no new emerging barriers and facilitators across three consecutive focus groups ^26^. Focus groups were conducted in-person at the participating hospitals, or when not feasible, via Teams. Each session was conducted by a moderator (LvV) and observer (SJ), both female doctors, conducting research in the field of neonatal infections, and LvV trained in qualitative interviewing. No relationship with the participants was established prior to the focus group; the observer and moderator introduced themselves, including their occupation and interest on improving EOS care. Afterwards, the moderator presented a 10-minute introduction of the three antibiotic stewardship interventions and started the discussion using a semi-structured interview guide (Supplemental file 2). Topics included perspectives on current early-onset sepsis care followed by the three antibiotic stewardship interventions. At the end of each session, the observer summarized the main findings, after which participants were invited to highlight any missed points or clarify misunderstandings. All focus groups were audio recorded after participants’ verbal consent and lasted approximately 90 minutes. Additionally, SJ took field notes during the focus group, while LvV recorded theirs immediately afterwards.

### Data analysis

A rapid CIFR rapid approach, a form of directed content analysis, was used for analysis. This method improves time efficiency and is suited for research that builds upon a framework ^27, 28^. Initially, EI, SJ, and LvV developed a codebook by selecting relevant CFIR constructs from the updated CFIR codebook template and replacing broad construct text with project specific definitions. Based on this, a matrix was constructed to organize focus group data. During the focus groups, primary analyst (SJ) took detailed notes, including quotations, and immediately coded these notes into the CFIR-based matrix, while indicating any areas that needed further detailing. The secondary analyst (LvV) then listened to audio recordings and reviewed the notes, expanding on the initial coding. The data analysis was an iterative process, allowing for ongoing refinement and improvement of the analysis throughout the research process. Weekly analyst meetings were conducted to align interpretations and maintain coding accuracy. Major determinants were defined as themes emerging in the majority of the focus groups, whereas minor determinants were defined as themes emerging in at least two focus groups.

## RESULTS

From January 2024 to July 2024, 11 focus groups interviews were conducted. A total of 81 healthcare providers participated (Supplemental file 3). The average number of participants was 7 (range 5-12). Ten focus groups were held on site, and one took place via Teams. Data saturation was reached after 9 focus groups.

### Determinants of EOS calculator, PCT-guided therapy and iv-oral switch therapy implementation

We identified 34 barriers and 20 facilitators regarding the implementation of the three interventions in neonatal care. Determinants relevant for all interventions are presented first, followed by specific barriers and facilitators for each intervention. Quotes illustrating the determinants, alongside their corresponding CFIR constructs are presented in Tables 2–5. Major determinants are indicated with a (*).

### General barriers and facilitators

#### Inner setting and individual characteristics

A topic that emerged in all focus group interviews was stakeholders’ dissatisfaction with the current national EOS guideline, due to high treatment numbers when following its recommendations and lack of clear guidance on discontinuing antibiotics. This has led some departments to deviate from the national guideline, and integrate the EOS calculator, PCT-guided therapy, or iv-oral switch therapy in their local protocols (Supplemental file 4). Other hospitals adopted different approaches, such as choosing observation over antibiotics when two maternal risk factors are present or maintained adherence to the national guideline. The primary motivation for adopting interventions outside the national guideline was the paediatrician teams’ emphasis on evidence-based care. They expressed the belief that the evidence supporting these interventions is stronger than that backing the current guideline’s recommendations, justifying its use as a substitute. Also, a strong local antibiotic stewardship programme and hospital’s vision focused on delivering family-centred, home-based care, were cited as key facilitators for reviewing national policies and making local changes. A major general barrier, for maternity nurses and midwifes, is the current working infrastructure and high workload, as implementation of the interventions would lead to shorter hospitalisation and earlier discharge home and might involve additional checks. However, paediatricians noted that early discharges represent only a relatively small proportion of neonates, suggesting a manageable impact, and emphasized that the responsibility for home-treatment remains with them, eliminating this potential burden.

#### Individual’s domain

In hospitals that adopted one or more interventions, paediatricians and nurses reported the positive effect of a clearly visible opinion leader, guiding colleagues through the latest evidence and translating it into clinical practice.

#### Outer setting domain

A major barrier to adopt novel antibiotic stewardship interventions was the perceived external pressure to adhere to the national or affiliated academic hospital’s policies, as deviating from this can lead to challenges from other hospitals or inefficient care, especially during patient transfers.

### EOS calculator

#### Intervention domain

The major decrease in antibiotic prescriptions is viewed as key benefit of EOS calculator implementation by all stakeholders, and its design is universally considered clear and practical. The calculator’s recommendations align more closely with the paediatrician’s ‘gut feeling’ than current guidelines. The expected uniformity in policy is also reported as an important advantage compared to current practice. A key barrier reported by microbiologists, paediatricians, and nurses is the current lack of evidence on EOS calculator’s safety in the Dutch population. Paediatricians and residents fear that higher antibiotic thresholds may result in delayed treatment of EOS, of which the impact is currently unknown. Also, the calculator’s lack of consideration of non-infectious symptoms and its inflexibility with missing data were reported as important barriers by paediatricians and residents. Some paediatricians noted uncertainty about the added value of the EOS calculator for clearly healthy or ill neonates.

#### Inner setting domain

Structurally delayed or incomplete communication of maternal information from the obstetric to neonatal department was reported as major barrier for EOS calculator use. Paediatricians, residents, and nurses also identified the incompatibility of the EOS calculator’s recommendations with Dutch workflows. The anticipated increase in neonates monitored in the maternity ward, instead of treated in the neonatology unit, is expected to result in capacity challenges.

#### Individuals’ domain

Midwives, maternity nurses, and neonatal nurses expressed the belief that maternal temperature should not be heavily weighted in the calculator’s risk assessments. Moreover, concerns were raised about the capability of obstetric nurses to provide frequent monitoring of neonates who are no longer receiving antibiotics under the new protocol.

#### Implementation process domain

Most paediatricians and residents considered electronic health record (EHR)-integration of the EOS calculator essential. Hospitals that already implemented the EOS calculator also reported improved communication between the obstetric and neonatal department by using an EHR-integrated standardized consultation.

### PCT-guided therapy

#### Intervention domain

Paediatricians reported a lack of evidence for higher-risk neonates as a barrier to using PCT, as research focuses on low-to medium-risk neonates. Moreover, they highlighted the conflict between the NeoPInS algorithm including an early CRP for risk categorisation and the low predictive value of post-partum CRP values in literature. PCT’s complexity due to the lack of a universal cut-off value and higher costs compared to CRP were also reported as challenges. Most stakeholders viewed the shorter exposure to treatment as an important facilitator, despite hospital management’s concerns about reduced revenue from earlier discharges in relation to the additional costs.

#### Inner setting domain

A key barrier to PCT-guided therapy in four hospitals was the lack of equipment to determine PCT. Moreover, according to nurses, the suggested timing of PCT blood draws may not align with current lab schedules. Nurses, paediatricians, and residents also reported concerns about the increased number of blood draws, contrasting their universal mission to minimize patients’ discomfort.

#### Individual’s domain

Paediatricians’ reliance on familiar CRP testing, alongside a lack of knowledge on interpreting PCT results, was reported as barrier. Moreover, they expressed hesitance to discontinue antibiotic treatment before receiving blood culture results, even when PCT values indicate its safety. Nurses questioned the relevance of reducing antibiotic duration by 24 hours. However, the need for a clear biomarker cut-off value, missing in current practice, serves as a motivator for paediatricians and residents to adopt PCT guided therapy.

### IV-to-oral switch therapy

#### Intervention domain

Microbiologists, paediatricians, and general practitioners were sceptical about the value of the evidence on IV-oral switch therapy, as they argued whether neonates with a negative blood culture should continue treatment at all. However, recognizing that in practice in some of the neonates with a negative culture treatment is continued regardless, the benefits of oral therapy at home are reported considerable by all stakeholders, including shorter neonatal discomfort from the IV, minimizing parent-child separation, and parents’ opportunity to receive maternity care at home.

#### Outer setting domain

Hospital pharmacists and microbiologists reported national medicine shortages affecting availability of amoxicillin suspension as barrier. Paediatricians, general practitioners, and pharmacists reported the pharmacy’s weekend fee for patients as barrier during weekends. Media coverage of the IV-to-oral switch therapy was reported to facilitate adoption.

#### Inner setting domain

There is a collaborative priority of family-centred care among all stakeholders, and early discharge home is regarded as a significant facilitator of this objective. However, communication practices from the hospital to maternity nurses at discharge were reported to be often inadequate or absent, hindering maternity nurses to provide informed support to parents. This communication issue is compounded by the fact that, unlike general practitioners and midwives, maternity nurses lack access to the health system’s digital communication platform. General practitioners and midwives emphasized the importance of receiving discharge letters including advice on surveillance. Moreover, maternity nurses reported the lack of access to knowledge sources regarding EOS as barrier for taking on extra responsibilities.

#### Individuals’ domain

Paediatricians look forward to IV-oral-switch therapy yet are hesitant to discharge neonates with markedly increased inflammatory markers. Low parental literacy or socio-economic status were reported as barrier for prescribing oral therapy at home. Furthermore, paediatricians expressed concerns on potential overuse of treatment, as the threshold to prescribe oral therapy may feel lower than prescribing IV therapy. This is compounded by paediatrician’s low trust in blood cultures, whether due to low sample volume, maternal antibiotics, or neonatal clinical status, resulting often in continuation of treatment as precaution in culture-negative cases.

#### Implementation process domain

Early engagement of the hospital’s pharmacy and microbiology department was reported to facilitate presence of resources and smooth roll-out of IV-to-oral switch therapy.

**Table 1.** Overarching determinants influencing implementation allocated to CFIR construct.

| CFIR Domain | CFIR Construct | Barrier/facilitator | Group | Quote |
| --- | --- | --- | --- | --- |
| Outer setting domain | Barriers |  |  |  |
|  | External pressure | *Pressure to follow national guideline, or guideline of academic hospital | P | *We stick to the academic hospital's protocol as much as possible, because no one wants to get reprimanded by the big brother.* |
| Inner setting domain | Barriers |  |  |  |
|  | Work infrastructure | *High current working pressure | MN, M | *When newborns are discharged home earlier, it will add to our workload. I assume we also need to perform extra checks. Honestly, we are already struggling to manage the workload as it is.* |
|  | Facilitators |  |  |  |
|  | Tension for change | *Dissatisfaction with current national guideline | P, PR, N, M, MN, GP, Mi, Ph | *After the national guideline was introduced a few years ago, we felt we were over-treating. That's why we've already adjusted the protocol. We no longer treat infants based on solely maternal risk factors. And we introduced PCT-guided therapy a few months ago.* |
|  | Learning-centeredness | Team's emphasis on evidence-based care | P, PR | *The driving force behind guidelines in this hospital is simply the best available evidence. Guidelines from nearby hospitals or the national guideline have less influence.* |
|  |  | Strong antibiotic stewardship program | P, PR, Mi, Ph | *There is much attention for antibiotic stewardship in our hospital. The A-team is constantly reminding us to focus on ways to improve antibiotic usage.* |
|  | Recipient centeredness | Hospital's focus on 'right care, right place', focusing on care at home | P, PR, N, Mi, Ph | *As colleagues, we feel really motivated to improve care because of the hospital's motto, 'right care, right place'.* |
| Individual | Facilitators |  |  |  |
| characteristics | Opinion leader | Clearly visible opinion leader, who is teaching colleagues the newest evidence | P, PR, N | We have two really enthusiastic colleagues who are on top of the newest evidence. They share their knowledge with the team which leads to discussing policy changes. |

*\*Indicates a key barrier, defined as reported during the majority of focus groups (>6 focus groups)*
*P=paediatrician, PR=paediatric resident, N=neonatology nurse, M=midwife, MN=maternity nurse,*
*Ph=pharmacist, Mi=microbiologist, GP=general practitioner*

**Table 2.**
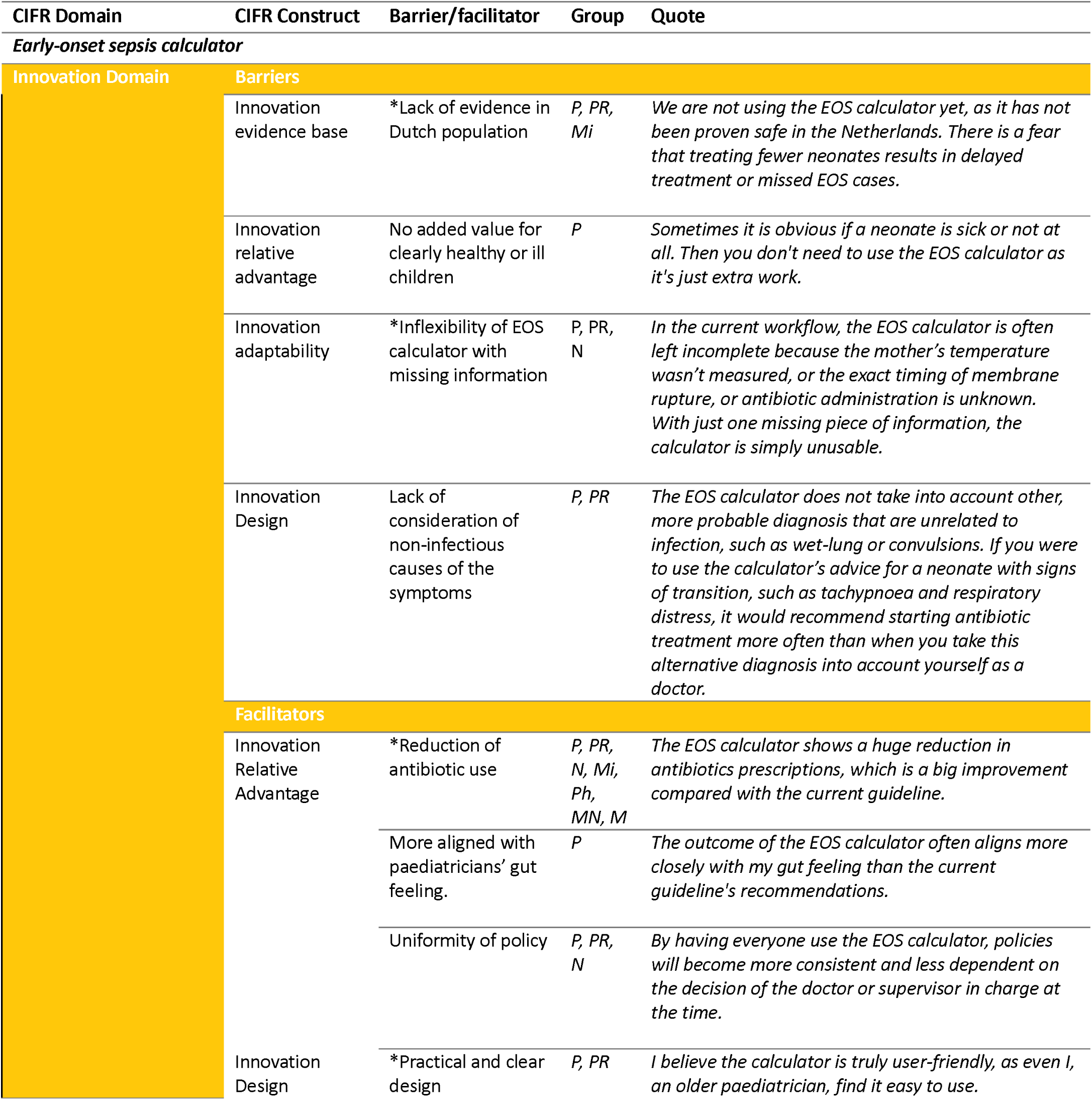
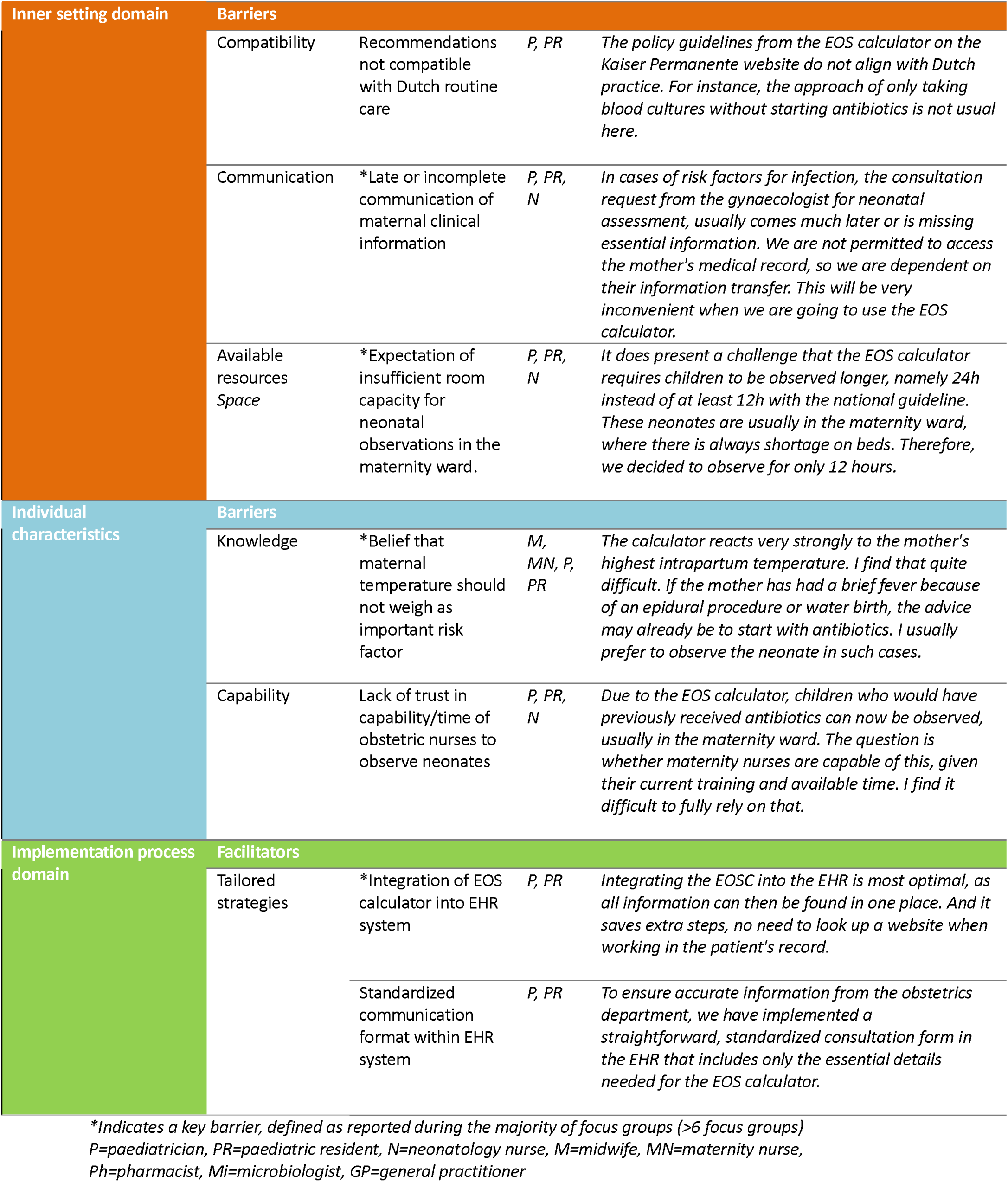
Determinants of EOS calculator implementation allocated to CFIR construct.

**Table 3.** Determinants of PCT-guided therapy implementation allocated to CFIR construct.

| CIFR Domain | CIFR Construct | Barrier/facilitator | Group | Quote |
| --- | --- | --- | --- | --- |
| **Procalcitonin-guided therapy** |  |  |  |  |
| **Innovation Domain** | **Barriers** |  |  |  |
|  | Innovation evidence base | Lack of evidence for high-risk neonates | *P, PR* | *The evidence from the NeoPInS study is not sufficient to guide treatment for all patients. It is unclear how to act in cases of higher infection risk. Our PCT-guided protocol is therefore partially based on the NeoPInS study's findings but also incorporates our own interpretation for the higher risk patients that the study does not address.* |
|  |  | *Algorithm in contrasts with literature on CRP predictive value | *P, PR* | *We do not determine CRP at the start of infection since it has no added value as clearly shown in the literature. It has low predictive value, as it has a wider normal range in the beginning of life. Therefore, I don't understand why this should be known to determine applicability of PCT-guided therapy.* |
|  | Innovation complexity | Complex use because of nomogram | *P, PR* | *Procalcitonin is more challenging to use than CRP due to the requirement of utilizing a nomogram. Clinicians must reference the normal values at different time points, as there isn't a single cut-off point, which adds complexity. Familiarizing oneself with this process takes time and practice.* |
|  | Innovation costs | *Additional laboratory costs | *P, M* | *Our hospital does not yet have resources to determine PCT, so if this needs to be obtained, it is an additional cost. PCT determination is also much more expensive than CRP. So, the added value must be very clear in order to consider it.* |
|  |  | Loss of income for the hospital | *HM* | *We have received some critical questions from the hospital management because there is less income when neonates can go home earlier, while we incur more costs on the PCT provisions.* |
|  | **Facilitators** |  |  |  |
|  | Innovation relative advantage | *Shorter harmful effects of treatment | *P, PR, Mi, Ph, GP* | *I think this will ensure that the total duration of antibiotics is shorter, which I think is particularly important with gentamicin. That stuff is really harmful.* |
| **Inner setting Domain** | **Barriers** |  |  |  |
|  | Available resources – equipment | *Unavailability of equipment for PCT testing | *P, PR, Mi* | *We don't have equipment for PCT determination available here. We can send it to a university hospital, but then it takes 24 hours until you get the result. There is no benefit from that.* |
|  | Compatibility | Insufficient compatibility with scheduled laboratory rounds | *N* | *In our hospital we have standard laboratory blood drawing rounds, which will not always correspond to the times when we should be drawing blood following the NeoPInS algorithm* |
|  | Mission alignment | *Conflicts with mission to improve | *P, PR, N* | *When using the NeoPInS algorithm, blood needs to be taken more often, which is an uncomfortable procedure* |
|  |  | patient comfort |  | *for the newborn. We always try to limit the blood draws as much as possible, so it is a bit of a dilemma to implement this.* |
| Individuals Domain | Barriers |  |  |  |
|  | Motivation<br>*Stage of change* | Tendency to hold on to familiar CRP testing | *P, PR* | *I believe many of my colleagues will struggle to adapt to using another biomarker. CRP is deeply ingrained in our routine, while PCT remains relatively unfamiliar.* |
|  | *Hesitance* | *Hesitancy to discontinue treatment before blood culture results are available | *P* | *We already use PCT-guided therapy here, but we typically wait for the blood culture results before making decisions. Most paediatricians are hesitant to stop treatment without having this information first, fearing a missed positive blood culture.* |
|  | Capability<br>*Knowledge* | Lack of knowledge on correct PCT-value interpretation | *P, PR* | *I am confident that many paediatricians will opt to continue a full course of antibiotics when PCT levels exceed the reference range, assuming it confirms infection. However, treatment can be safely discontinued if the blood culture is negative, and the newborn is clinically stable. This presents a significant pitfall, as there is a tendency to place too much emphasis on elevated biomarkers.* |
|  |  | Lack of knowledge on benefits of shorter treatment duration | *N* | *Does it really matter if you treat for 24 hours instead of 48 hours? Both are quite short. What is the added value?* |
|  | Facilitators |  |  |  |
|  | Need | *Need for clear biomarker cut-off | *P, PR* | *We need clear biomarker cut-offs to stop AB in the case of a negative blood culture. There is a lot of variation in what CRP cut-offs are currently used. PCT-guided may be a nice solution to get more uniform policy.* |

*\*Indicates a key barrier, defined as reported during the majority of focus groups (>6 focus groups)* *P=paediatrician, PR=paediatric resident, N=neonatology nurse, M=midwife, MN=maternity nurse, Ph=pharmacist, Mi=microbiologist, GP=general practitioner*

**Table 4.**
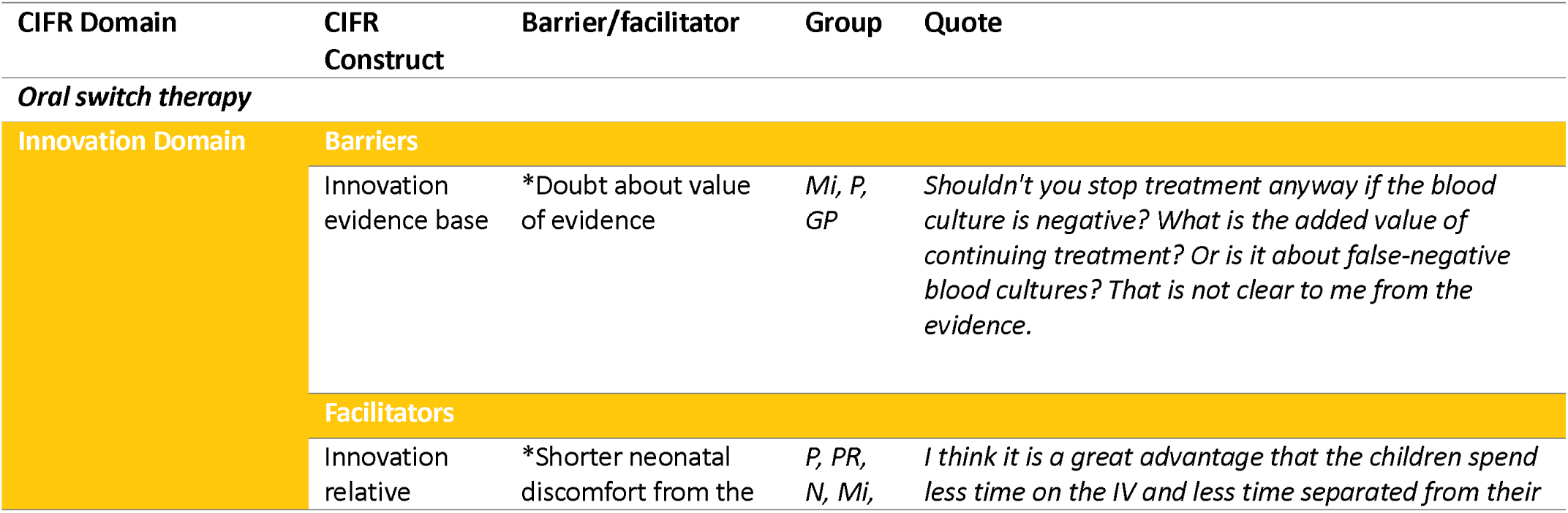
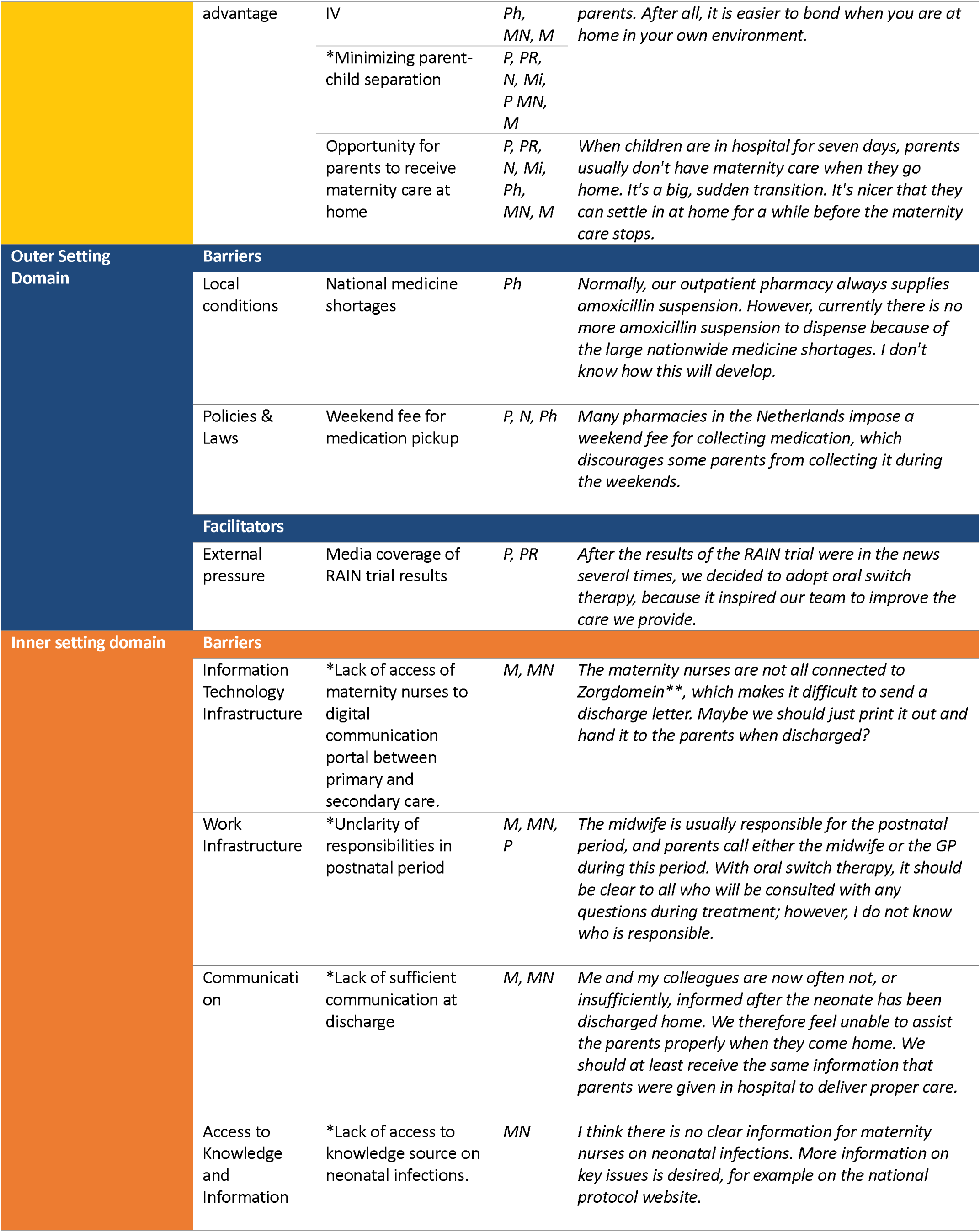
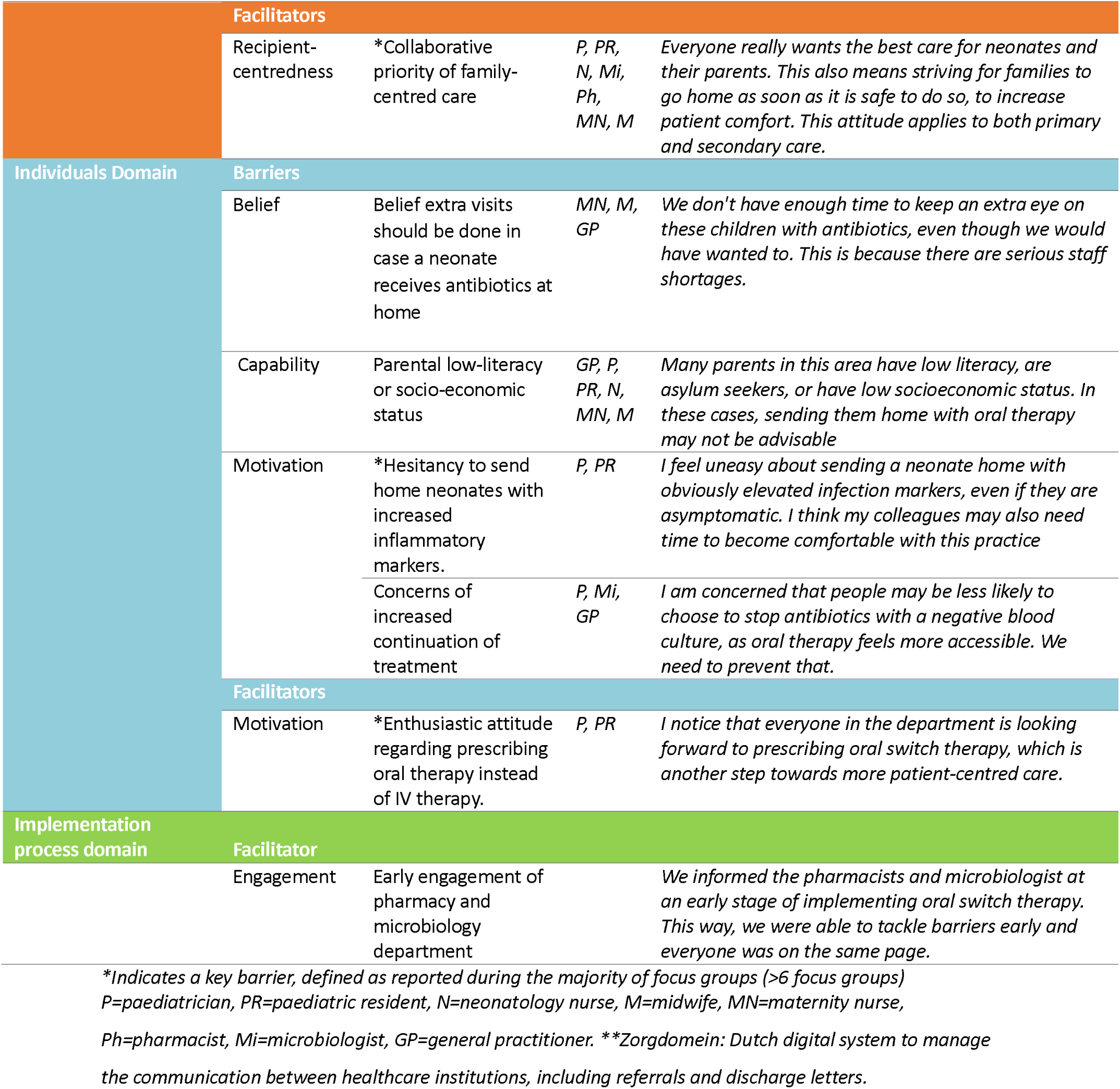
Determinants of oral switch therapy implementation allocated to CFIR construct.

## DISCUSSION

We systematically evaluated the determinants affecting the implementation of three antibiotic stewardship interventions in neonatal care. The overlapping recurring barriers were found in the following domains: inner setting, intervention characteristics, individuals, and a few in the outer setting. The positive values and barriers can be consolidated in the following themes: balancing evidence with professional core values, managing care shifts and enhancing communication, and conflicts with un-updated national guidelines.

### Balancing evidence with professional core values

Even though a great facilitator for all interventions is the evidence for reducing antibiotic treatment and promoting family centred care, at the same time the lack of specific evidence seems a barrier for implementation. On one hand, there is reluctance to deviate from guidelines and apply evidence-based strategies, especially if they are not based on randomised controlled trials (RCTs). On the other hand, stakeholders are willing to diverge from guidelines that are not supported by the newest evidence. This is in particular the case for the EOS calculator, where the lack of high-level evidence within specifically the Dutch population is the greatest barrier, as it leaves uncertainty for safe usage amongst stakeholders. It is known that clinical risks lead to defensive medicine, which might ask for higher level of evidence when disastrous outcomes are at stake ^29^. The outcomes of a RCT comparing the safety of the EOS calculator to the Dutch national guideline may address this hurdle ^30^. However, though high-level evidence is essential, it does not automatically lead to successful implementation or guideline adherence, as controlled settings differ from the real-world application ^15^. The evidence concern for the PCT-guided therapy is the usage of CRP for risk classification, while CRP values in healthy neonates vary physiologically postpartum and thus have low predictive value ^31^. For the IV-to-oral switch therapy, some were sceptical about the value of evidence and argued whether a neonate with a negative blood-culture should continue treatment at all. However, low confidence in neonatal blood cultures that exists mainly among paediatricians, results in the common practice to treat culture-negative cases. In accordance with literature, the low confidence was found to be attributed to the limited volume of blood collected, concerns that maternal intrapartum antibiotic use may lead to false-negative results, and cases where the neonate’s clinical status does not match the culture results ^32^.

Family-centred care is a well-known concept within paediatric and neonatal healthcare as it has positive effects on the neonate and family such as bonding and neurodevelopmental outcomes ^33, 34^. It includes respect and collaboration, information sharing, support, flexibility in care, and promoting family well-being ^33–35^. The IV-to-oral switch resonates particularly well with this concept as it shortens neonatal discomfort from IV, minimizes parent-child separation, and gives parents’ the opportunity to receive maternity care at home. Since a shared concern of patient-centred care is found to be an important facilitator for collaboration between healthcare workers, emphasis on this should be placed when informing and motivating stakeholders ^18^.

### Managing care shifts and enhancing communication

Implementing all interventions shifts the delivery of care among healthcare providers, initiating resistance to change among those the shift is aimed at. This inner setting barrier was greatest for clinical and primary care midwives and maternity nurses, as this shift would create: a higher workload, reduced admission capacity, and introduce new responsibilities in caring for neonates with higher sepsis risk, being commonly reported barriers for change and expanding roles ^36–39^. This underscores the relevance analysing the consequences of novel interventions for all involved healthcare workers, even if they are less visible, to get a realistic idea of the shift in workload. Secondarily, to overcome these barriers, clear agreements should be made on the responsibility in care for neonates, especially in the primary care setting, as lack of role clarity hinders role expansion ^40^. Moreover, empowering their knowledge base on EOS physiology, including neonatal symptoms of infection and instruction on oral antibiotic administration, is needed to support their new roles ^40^.

Furthermore, insufficient communication within the hospital walls and between primary and secondary care, raised awareness that improvement is needed for safe implementation. Timely, frequent, and consistent information sharing are essential for interprofessional communication, especially for the EOS calculator, as its functionality is dependent on the complete input from both departments. To achieve proper collaboration, there should be awareness and shared knowledge on the EOS calculator among departments ^41, 42^. Moreover, integration of the tool into an EHR could reduce workload and may promote communication efficiency, accuracy, and saves time ^43, 44^. For the PCT-guided therapy, aiming for mutual knowledge and understanding trough a proper business case, presented by a local implementation champion, is needed to convince management for in hospital PCT-testing, as most regional hospitals reported lack of equipment and concerns about additional costs although evidence shows the cost-effectiveness of PCT-testing in low-risk neonates from a public health perspective ^41, 45–47^. In case of oral switch therapy, attention should be given to timely communication at discharge from hospital to primary care maternity nurses and midwives. Shared knowledge and collective awareness of the neonates’ status are important elements of safe communication ^42^. Standardized discharge letters and facilitating maternity nurses’ access to the national digital communication platform are steps to improve communication practices ^48, 49^.

### Conflicts with un-updated national guidelines

Healthcare professionals unanimously expressed dissatisfaction with the current EOS guideline due to the high number of antibiotic prescriptions and unclarity of criteria to discontinue treatment. Un-updated guidelines may raise concern in healthcare professionals which could lead to distrust, causing clinicians to either implement other evidence themselves or deviate from the guideline based on clinical experience ^50^. This highlights the importance of frequently updating guidelines with the newest evidence. The ‘living guidelines’ approach, a dynamic set of recommendations that facilitate continuous guideline revisions, could be a solution to this ^51^. This entails a collaborated team that systematically and rapidly monitors new evidence, prioritizes which recommendations in the guideline need updating, and effectively communicates via channels to policymakers, clinicians and other stakeholders ^51^. Even though digitalization and emerging software could facilitate this ^51^, a process consisting of dissemination, implementation, and best practice recommendations are required ^52^. When national guidelines lag behind in new evidence, it is crucial for hospitals to proactively update their local protocol to avoid uninformed practices ^50^. The key driving factor for this accomplishment is an opinion leader, who places the best available evidence above the national guidelines and guides a team through the implementation process ^53^. Moreover, a climate focused on learning and patient centeredness, along with a clear vision of “providing the right care in the right place”, and a strong local antibiotic stewardship were key for critically reviewing local policies and making changes. All three concepts are considered motivators in current literature ^54–56^. However, since these are hospital-specific, strategies to promote cross-hospital exchange should be pursued. Besides sharing experiences and knowledge, benchmarking, involving data sharing in standardized ways to compare outcomes and processes between hospitals, is critical for creating transparency, collaborative learning, and improving clinical decision-making, ultimately leading to improvements in neonatal antibiotic stewardship ^4^.

### Strengths and limitations

This is the first study to systematically analyse determinants of implementing three antibiotic stewardship interventions in neonatal care. By analysing the EOS treatment process from initiation to discontinuation or alteration of antibiotics, it facilitates comprehensive implementation into the national guideline. Furthermore, we reached data saturation by performing multiple focus group in various regions, including all medical professionals involved in the entire treatment sequence, enabling different perspectives. Some barriers identified were time-sensitive, such as a temporary national shortage of amoxicillin suspension, which may be ignored in implementation strategies. Conversely, other time-sensitive barriers in the future might not be foreseen with in this study. Another limitation to the study is that participation was voluntary, which could increase selection bias towards urge for change, perhaps shifting attitudes towards interventions positively. Moreover, the member check was performed after each group discussion, as opposed to mailing members. This practical approach may have increased responses but can lead to withheld beliefs in a group setting.

## CONCLUSION

We identified 34 barriers and 20 facilitators in the implementation of three neonatal antibiotic stewardship interventions, including the EOS calculator, PCT-guided therapy and IV-to-oral switch therapy. These barriers and facilitators can be consolidated in the following themes: balancing quality of evidence with professional core values, managing care shifts and enhancing interdisciplinary communication, and conflicts with un-updated guidelines. The next step is to develop evidence-based implementation strategies that incorporate address these factors.

## Data Availability

The data supporting the findings of this study are not publicly available and will not be shared due to the personal and sensitive nature of the data collected from focus group participants.

## ACKNOWLEDGEMENT

The authors would like to express their gratitude to all the hospitals participating in the Protect-Neo study for their contributions.

