## Supplemental file 4 for "Facilitators and barriers when implementing antibiotic stewardship interventions in neonates at risk of early-onset sepsis"

**Supplementary Table S2**. Adoption status and study participation

| | **Implemented intervention as regular care at time of focus group discussion (yes/no)** | | |
|---|---|---|---|
| **Hospital** | *EOS-calculator* | *PCT-guided therapy* | *IV-to-oral switch therapy* |
| *A* | No | No | No |
| *B* | No | No | No |
| *C* | Yes | Yes | Yes |
| *D* | No | Yes | No |
| *E* | No | No | No |
| *F* | Yes | No | Yes |
| *G* | No | No | Yes** |
| *H* | Yes | Yes* | No |
| *I* | No | No | Yes |
| *J* | No | No | No |
| *K* | Yes | No | No |

*Grey boxes display hospitals' participation in the EOS calculator RCT, NeoPInS study or RAIN study.*

**Hospital uses adapted version of PCT-guided therapy. Adaptations:*

*- PCT is measured in all neonates, not only in low and intermediate risk cases.*

*- If PCT falls within normal range at t=12 and t=24, blood culture is still awaited before decision to discontinue antibiotic course.*

***Hospital was already applying oral switch therapy before the publication of the RAIN study. They switch to oral therapy after 5 days of intravenous therapy if the neonate is in good clinical condition and has a negative blood culture.*
