## Supplemental file 3 for "Facilitators and barriers when implementing antibiotic stewardship interventions in neonates at risk of early-onset sepsis"

**Table S1.** Characteristics of focus groups participants

| | **Age (Years)** | **Working experience (Years)** |
|---|---|---|
| | *Median (IQR range)* | *Median (range)* |
| Paediatricians (n=15) | 45 (35-63) | 13 (3-30) |
| Residents/physician assistants (n=8) | 28.5(26-57) | 2.5(0.5-26) |
| Neonatal nurses (n=14) | 46 (25-59) | 15 (3-26) |
| Pharmacists (n=10) | 43 (35-47) | 11 (5-20) |
| Microbiologists (n=8) | 38.5 (33-50) | 5 (1-21) |
| Primary care obstetricians (n=10) | 41 (25-62) | 14 (2,5-22) |
| Primary care maternity workers (n=10) | 47 (41-55) | 13 (3-33) |
| General practitioners (n=6) | 47 (36-65) | 15 (12-29) |
