## Supplemental file 2 for "Facilitators and barriers when implementing antibiotic stewardship interventions in neonates at risk of early-onset sepsis"

**Interview guide**

*Background information*

A brief presentation will be given by the researcher (LvV) discussing the following:

- Introduction of researchers (job, background, relationship to the research topic)
- Exact purpose of the focus group (obtaining perspectives on current EOS care and possible implementation of new interventions).
- Background information on:
    - The EOS calculator
    - PCT-guided therapy
    - IV-Oral switch therapy

Introduction round

Can you introduce yourselves? Please name the following points:

- Your name
- Your profession
- In what way you deal with neonates at increased risk of infection in your work
- From the perspective of your profession: your most important core value in caring for neonates at risk of infection

Current care & collaboration

- How do you currently jointly manage the care of neonates at risk of infection in the *hospital* and surrounding primary care network?
    - What are everyone's responsibilities?
    - In what way are decisions made about changes to local protocols?
    - How is change in policy handled in your network, e.g. implementation of new strategies?

Antibiotic initiation/EOS calculator

- How do you view the current policy recommendations regarding starting antibiotics in neonates at risk of EOS?
- Is the EOS calculator already applied in your hospital/region, or do you already use the EOS calculator?
    - If yes:
        - Why was it chosen/why did you choose to use the calculator?
        - How do you experience using the calculator?
        - Are there any issues you encountered when using the calculator?
        - What promotes the use of the calculator?
    - If no:
        - Why did you choose to not use the calculator?
        - Would you like to use the calculator in the future?

            - If no: what stops you from doing so?
- What is needed to make use of the EOS calculator a success in your hospital and region?
- The EOS calculator was designed in the United States. How important is it for you to test and/or further develop this calculator specifically for the Dutch population?

<u>Discontinuation of antibiotics /PCT-guided therapy</u>

1. How do you view the current policy recommendations regarding the decision to stop antibiotics in a neonate who is suspected of EOS?
2. Is PCT-guided therapy already used in your hospital/region, or do you already use PCT-directed stopping?
    1. If yes:
        - Why was it chosen/why did you choose to use PCT-guided therapy?
        - How do you experience this?
        - Are there any issues you encountered while using PCT-directed cessation?
        - What promotes the use of PCT-controlled cessation?
    2. If no:
        - Why do you not use PCT-guided therapy?
        - Would you like to use PCT-guided therapy in the future?
            - If no: what stops you from doing this?
3. What is needed to make PCT-guided therapy a success in your hospital and region?

<u>Continuation of antibiotic treatment/oral switch therapy</u>

1. How do you view the current policy recommendation regarding treatment continuation in case of a negative culture and good clinical status?
2. Is oral switch therapy already applied in your hospital/region, or do you already apply oral switch therapy?
    a. If yes:
        - Why was it chosen/why did you choose to apply oral switch therapy?
        - How do you experience this?
        - Are there any issues you encounter when applying oral switch therapy?
        - What promotes the application of oral switch therapy?
    b. If no:
        - Why is oral switch therapy not applied?
        - Would you like to apply oral switch therapy in the future?
            - If no: what stops you from doing so?
3. What is needed to make application of oral switch therapy a success in your hospital and region?
